# Easing social distancing index after COVID-19 pandemic

**DOI:** 10.1101/2020.06.11.20128165

**Authors:** Sam Li-Sheng Chen, Amy Ming-Fang Yen, Chao-Chih Lai, Chen-Yang Hsu, Hsiu-Hsi Chen

## Abstract

**Context:** Easing social distancing (ESD) is a global public health issue in post-pandemic period of COVID-19 and requires a simple index for real time assessment.

**Objective:** We aimed to develop a simple index for ESD to quantify the impacts of social distancing for reducing confirmed infected cases, optimal triage and care of patients for recovery, and critical care capacity for reducing death from COVID-19.

**Design, Setting, and Participants:** Data on the retrospective cohort of 185 countries with reported numbers on confirmed cases, recovery, and death from COVID-19 were retrieved from publicity available repository. Up to May 31, a total of 5,844,136 confirmed cases, 2,639,961 recovered, and 327,487 deaths were reported globally.

**Main Outcome Measures:** The ESD index measured by cumulative number of COVID-19 cases and recovery and case-fatality rate.

**Results:** We developed a simple index for the guidance of easing social distancing (ESD). If the ESD index is less than 1, ESD would be considered. The global ESD index declined from 3.87 at peak in March to 1.35 by the end of May, consisting of 56.76% countries/regions (105/185) with the ESD lower than one.

**Conclusion and Relevance:** This simple ESD index provides a quantitative assessment on whether and when to ease social distancing from local to global community.

## Introduction

While border controls and social distancing have been executed since the beginning of the Coronavirus disease 2019 (COVID-19) pandemic^1-4^ it is time to ponder over how to ease social distancing in the post-pandemic period with real time assessment ^5^ as reviving economic business and normal social activities have been urgently needed.

Though there are six criteria for countries while considering de-escalation by reversing restrictions or lockdown^6^, it is still unclear whether and when to implement the reopening policy. It is insufficient to merely consider the force of the spread of COVID-19.^2-4,7-11^ One has to consider the optimal allocation of COVID-19 with an efficient triage system to accelerate the rate of recovery from hospitalization or self-isolation and also take into account critical care capacity to avert death from COVID-19. To quantify the impacts of these three factors, a simple ESD index was developed for health decision-makers to do real-time assessment of COVID-19 at global, country, region, and community level.

## Material and Methods

The data for analysis were derived from the web-based real-time GitHub repository created by the Center for Systems Science and Engineering (CSSE) at Johns Hopkins University.^12^ CSSE operates daily updates upon publicly available data, including confirmed cases, recovered cases, and deaths from multiple sources. A total of 188 countries have reported confirmed COVID-19 cases (including presumptive positive cases and probable cases) at country and region level that is aligned with WHO situation reports.^13^ A total of 185 affected countries and regions were available from Jan. 22 till May 31, 2020 after excluding three countries or regions without information on recovery. Data on New York State were abstracted from GitHub repository.^14^

An index of easing social distancing (ESD) is expressed as follows.

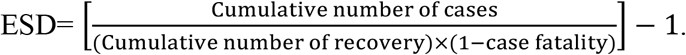

ESD is the ratio of cumulative confirmed cases to cumulative recovered patients without dying from COVID-19 that is captured by (1-case-fatality (f)) minus one during a fixed time period. The reason for subtracting one is that in an ideal scenario the aforementioned ratio would reach 1 when all confirmed cases have been recovered without death (case-fatality rate=0) and therefore ESD would approach zero, suggesting the region has a full recovery after the outbreak of COVID-19 and may return to the normal status.

However, it is impracticable to ease social distancing until the value of ESD reaches to 0. One has to consider the balance between the spread of COVID-19, the rate of recovery, and critical care capacity. The first element is to capture the information on the force in relation to the spread of COVID-19 after the implementation of social distancing that is often modelled by reproductive number. The second element is dependent on whether health care systems have the capacity of offering hospitalization based on efficient triage of COVID-19 patients. The third one is determined by critical care capacity that can stop the progression from acute respiratory distress syndrome (ARDS) to subsequent deaths. If the value of ESD index is larger than 1 it is still necessary to maintain social distancing because the rate of the spread of COVID-19 still outweighs the affordable capacity of hospitalization and critical care. If it is lower than 1 easing social distancing (ESD) can be considered. The degree of ESD lower than 1 is assessed by the inverse of the decile of ESD index form the lowest percentile (0.1) to the highest percentile (1). The multinomial distribution using the reported number of confirmed cases, recovery, and death in conjunction with Bayesian Markov Chain Monte Carlo method was used for the derivation of 95% credible interval (CI) for the ESD index.

## Results

### The ESD index for countries/regions worldwide

As of 31^st^ May, 5,844,136 confirmed cases, 2,639,961 recovered, and 327,487 deaths from COVID-19 were reported globally, which gave a case-fatality rate of 5.6%. The global ESD index was 1.345 (95% CI: 1.343-1.347), which suggests that easing social distancing has not been reached yet at global level. Namely, the spread of COVID-19 from hotspot to hotspot still exceeded the capacity of hospitalization and critical care for COVID-19 cases at global level.

**Figure 1** shows the temporal trend of global ESD index. The global ESD index began from the 1.141 (95% CI: 1.127-1.157) on Mar. 1 just before the announcement of COVID-19 pandemic and peaked on Mar. 29 (3.870, 95% CI: 3.848-3.892).

**Figure 1.**
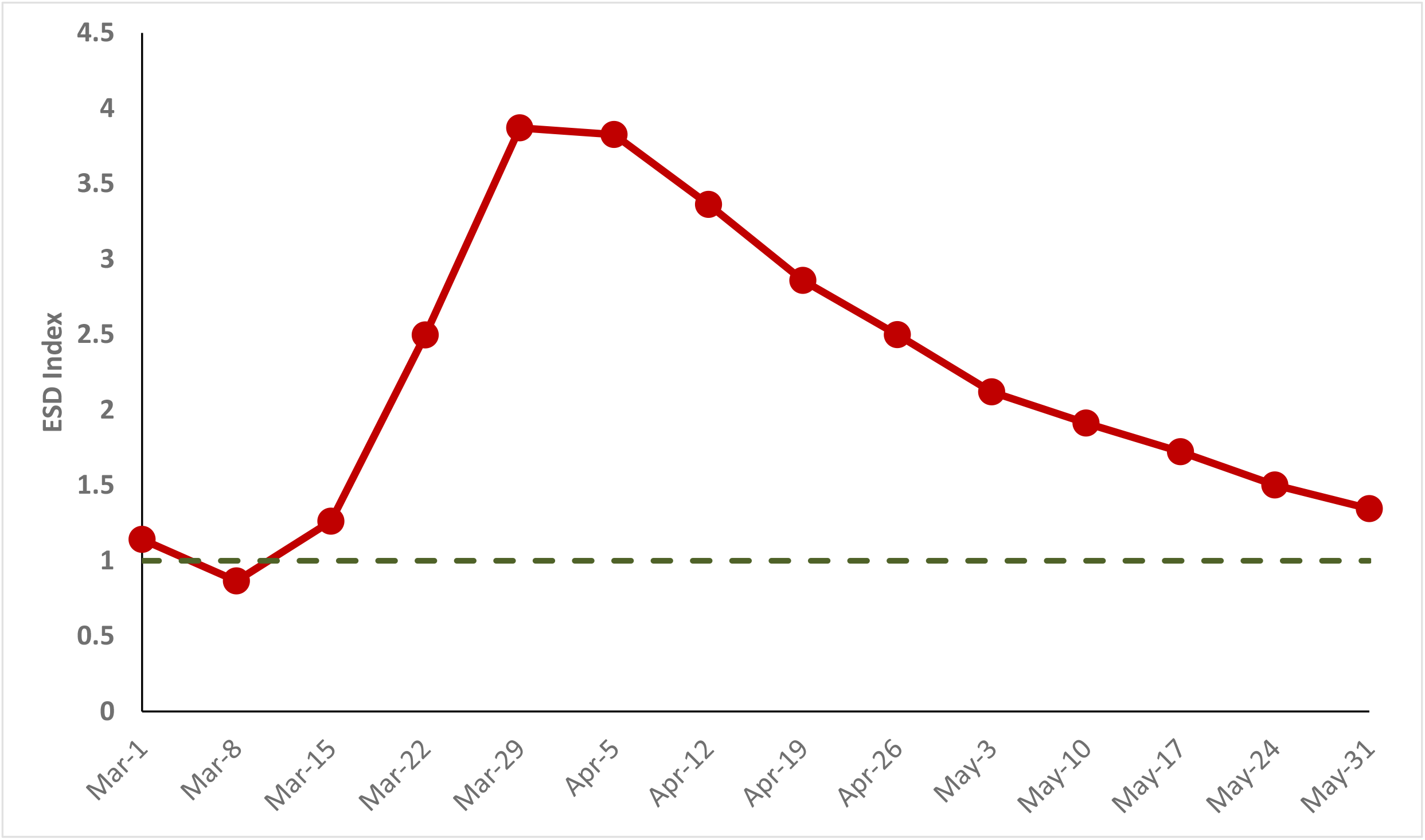
Temporal trend of global ESD index. ESD: easing social distancing

**Figure 2** shows the distribution of ESD index for 185 countries and regions. Countries with the color in red are less likely to ease social distancing whereas those with the color in green are the opposite. Of 185 countries/region, 56.75% (n=105) of the ESD index was smaller than 1.

**Figure 2.**
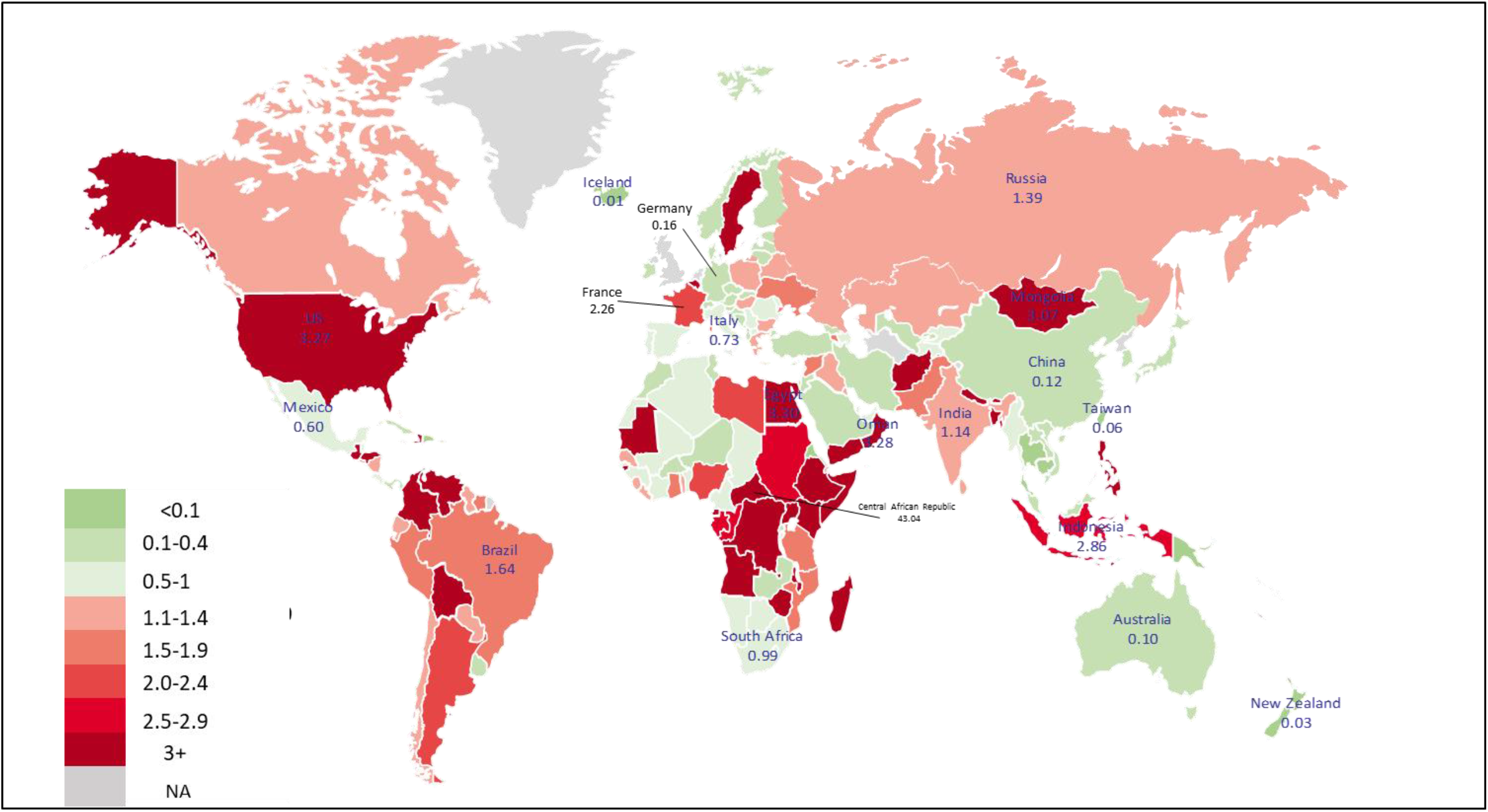
Global distribution of ESD index by the end of May. ESD: easing social distancing

According to the decile of the ESD index for each of two categories (≥ 1 and <1), **Figure 3** shows the frequencies on the index of ESD for the countries/regions globally aggregated by three groups for the ESD ≤ 1, including < 0.1 (n=16), 0.1-0.4 (n=52), 0.5-1 (n=37), and by five groups group for the ESD >1, including, 1.1-1.4 (n=26), 1.5-1.9 (n=10), 2-2.4 (n=4), 2.5-2.9 (n=5) and 3+ (n=35).

**Figure 3.**
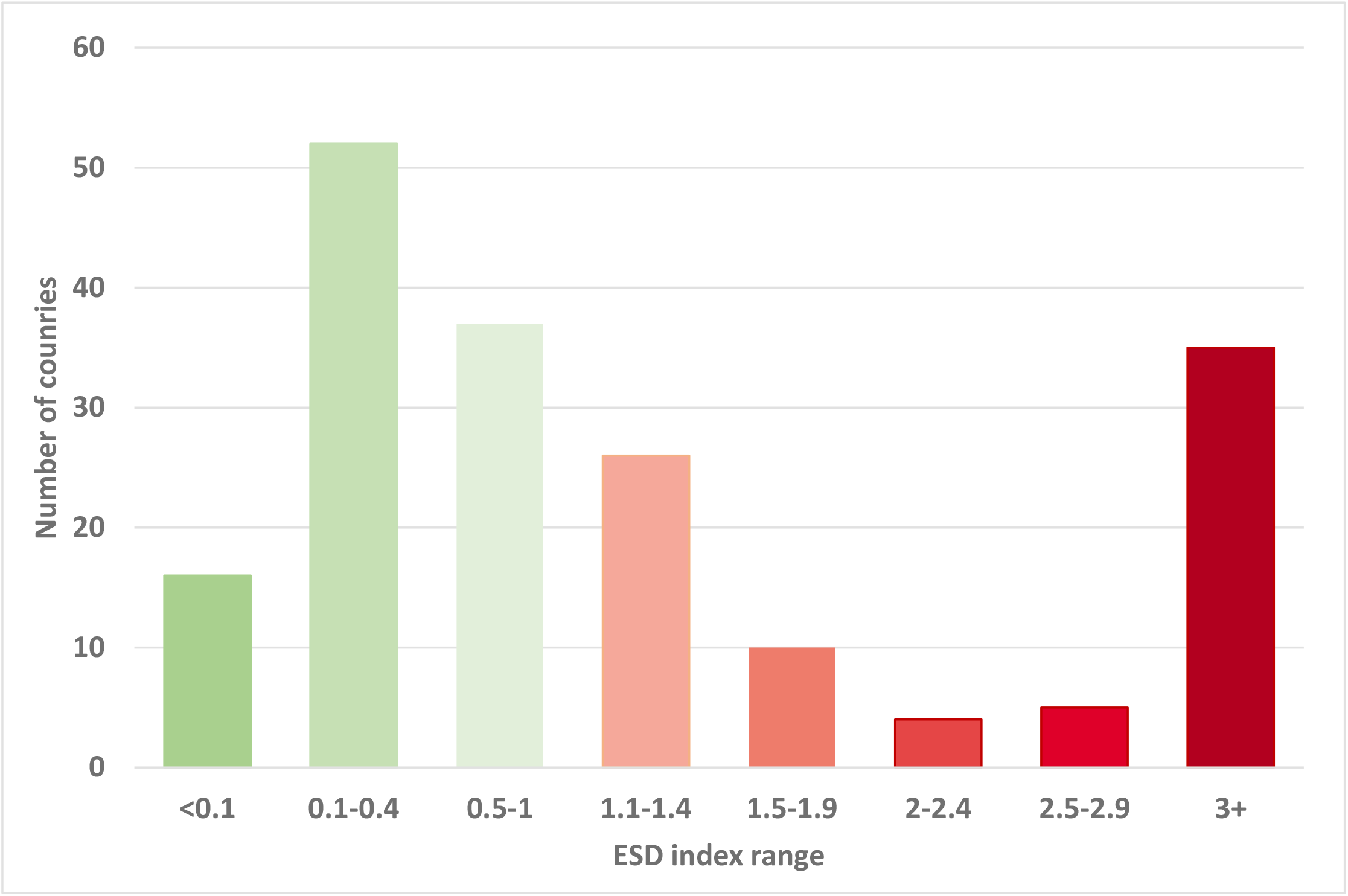
Number of countries/regions by ranges of ESD index by the end of May. ESD: easing social distancing

### The ESD index used for different scenarios

The ESD index can be adapted to different kinds of scenarios featuring with differential influences of three elements.

Countries and regions show a low ESD index such as Iceland and Taiwan had high recovery rate (Iceland: 99%, Taiwan: 96%) and low case-fatality (Iceland: 0.6%, Taiwan: 1.6%), both indicate that the spread of COVID-19 has been contained and both the patient triage system and critical care capacity were affordable to meet the needs for COVID-19 cases during the pandemic. For countries such as Germany (ESD index: 0.163; 95% CI: 0.160-0.166), a moderate case-fatality rate (4.7%) was compensated by the high recovery rate (90%) through early detection followed by efficient patient triage and high capacity of hospitalization.

Countries with high ESD index such as France (ESD: 2.257; 95% CI: 2.235 - 2.278) were attributed to high case-fatality rate mainly resulting from insufficient capacity of critical care. The high ESD index was, to a greater extent, due to 15.2% case-fatality rate (around 2.7 times the average worldwide) and, to a lesser extent, due to modest recovery rate (36.2%, around 0.8 of the average worldwide).

The impact of high transmission of SARS-COV-2 and low recovery rate (17.8%) would result in extremely higher ESD demonstrated by New York State (ESD: 5.011; 95% CI: 4.967-5.053) even though the case-fatality rate (6.4%) was comparable to the average worldwide.

### Resurgence of COVID-19 epidemic in local community

The ESD index can be also applied to evaluating whether social distancing has to be re-executed. Using South Korea as an example, although the overall ESD till the end of April was 0.22, the ESD indices from May 1 up to May 16 and May 31 were estimated as 2.39 and 4.93, respectively, indicating that the outbreak was re-emerging and might call for re-strengthening social distancing measures.

## Conclusion

In summary, a simple index for easing social distancing was developed to aid health policy-makers in the assessment of whether and when to ease social distancing in post-pandemic period from local to global community.

## Data Availability

All data are available from the authors.

## Authors’ contributions

SLSC, AMFY, and HHC conceptualize and design the framework of analysis. AMFY, CCL, and CYH abstracted and prepared the data for analysis. SLSC, AMFY, CYH, and HHC developed the methods. All authors agreed the findings and provided input on the revision of the manuscript.

## Data Sharing Statement

All data and codes for analysis are available from authors.

